# Knowledge and Practice of Breast Self-Examination and Clinical Breast Examination among Women in the Buea Health District, Cameroon: A Community-Based Cross-Sectional Study

**DOI:** 10.64898/2026.09.18.26363456

**Authors:** Ruth Tabi Amin, Arole Darwin Touko, Tadolonkeu Raissa Manemik, Notaya Diane Tazoho, Bassah Nahyeni

**Author notes:** **Corresponding author:** Arole Darwin Touko.

## Abstract

**Background:** Breast cancer is the most frequently diagnosed cancer and leading cause of cancer mortality worldwide, accounting for 2.3 million new cases and 685,000 deaths in 2020. In Cameroon, breast cancer is the leading cancer among women, representing 34.1% of new female cancer cases. Where mammography is unaffordable and inaccessible, BSE and CBE may support early detection through breast awareness; however, uptake remains poorly characterized in Cameroon.

**Methods:** A community-based cross-sectional study was conducted between June and December 2020 among 346 women aged 21–60 years across five health areas of the Buea Health District using probability-proportionate-to-size sampling. Data were collected via a structured questionnaire covering breast cancer knowledge and BSE/CBE knowledge and practice, scored as good (≥70%) or poor (<70%). Descriptive statistics, Chi-square tests, and multivariate binary logistic regression were performed using IBM SPSS Statistics version 21.0 (p < 0.05). Ethical approval was granted by the Ethics Committee of the Faculty of Health Sciences, University of Buea (reference number: 1199-04).

**Results:** The mean participant age was 28.3 years (SD = 7.0 years); most were single (67.0%) and university-educated (62.1%). Good breast cancer knowledge was demonstrated by only 24.0% (95% CI: 19.6%–28.4%), and good BSE/CBE knowledge by 26.9% (95% CI: 22.3%– 31.6%). Satisfactory BSE/CBE practice was observed in 22.3% (95% CI: 17.9%–26.7%). University education was independently associated with satisfactory BSE/CBE practice (AOR = 2.148; 95% CI: 1.544–2.649; p < 0.001). A significant positive association was observed between knowledge and practice (χ^2^ = 73.324, p < 0.001, φ = 0.460).

**Conclusion:** Knowledge and practice of BSE and CBE are critically low among women in the Buea Health District. University education is a significant independent predictor of satisfactory BSE/CBE practice. Urgent, multilevel community-based breast cancer education interventions particularly targeting women with limited formal education are needed to improve early detection and reduce breast cancer mortality in Cameroon.

## Background

Breast cancer is currently the most frequently diagnosed malignancy and the leading cause of cancer-related mortality among women globally [1]. In 2020, an estimated 2.3 million new cases and 685,000 deaths were recorded worldwide, representing a global burden of unprecedented magnitude [2]. While incidence rates have historically been higher in high-income countries, mortality rates are disproportionately elevated in low- and middle-income countries (LMICs), where women younger than 50 years account for a significantly greater proportion of deaths [3]. This disparity is attributable to delayed diagnosis, limited access to screening technologies, and the high cost of treatment [4,5]. The social and economic consequences of breast cancer in LMICs are profound, affecting not only individuals but also household welfare, childcare, and broader economic productivity [6].

In Cameroon, breast cancer is the most incident cancer across both sexes and accounts for approximately 16% of all cancer deaths [7]. Of the approximately 12,235 new cancer cases diagnosed among women in Cameroon in 2020, 34.1% (n = 4,170) were attributable to breast cancer [8] a marked increase from the 2,625 cases per 100,000 women recorded in 2012 [9]. This rising burden underscores the urgency of developing effective, context-appropriate early detection strategies.It is important to note that data collection for this study was conducted between June and December 2020, a period coinciding with both the COVID-19 pandemic and an escalation of the Cameroon Anglophone Crisis. Both may have influenced health-seeking behaviour, willingness to participate, and access to clinical services among study participants contextual factors that should be considered when interpreting these findings.

Early detection through screening and prompt treatment are the cornerstones of effective breast cancer control [2,4]. However, the majority of breast cancer cases in LMICs including Cameroon are diagnosed at advanced stages, largely due to inadequate access to diagnostic infrastructure [10]. Mammography remains the most validated screening modality, though major international guidelines, including WHO and USPSTF do not recommend population-based BSE programmes due to insufficient evidence of mortality reduction; instead, they endorse breast awareness as a means of facilitating early presentation [11–13]. CBE may complement mammography in low-resource settings, though evidence for its stand-alone efficacy remains limited [10]. Understanding women’s knowledge of and engagement with these approaches in Cameroon remains an important public health priority.

The theoretical basis of this study is grounded in the Health Belief Model (HBM) [14], which posits that health-related behavior such as BSE practice is influenced by an individual’s perceived susceptibility, perceived severity, perceived benefits, perceived barriers, and cues to action. This framework is used to contextualise rather than empirically test the patterns of knowledge and practice observed, as HBM constructs were not directly measured in this study.

Several studies conducted in the South West Region of Cameroon have explored BSE knowledge [12,15,16]; however, women’s awareness of CBE and their actual practice of both modalities remain poorly understood, particularly in urban communities. Importantly, the most recent comparable study [12] was conducted in a rural setting; an evidence gap exists for urban populations, which may have different knowledge profiles due to greater access to education and media. Understanding breast cancer knowledge and screening practices in the urban Buea Health District which hosts the University of Buea and harbours a comparatively educated population is therefore essential for developing targeted, effective educational interventions. The inclusion of women aged 21–30 years who constituted the majority of participants (73.1%) was deliberate: while this age group falls below the mean diagnostic age for breast cancer in Africa (30.6–60.8 years) [17], early knowledge acquisition and practice formation in young adult women are critical to establishing lifelong health behaviours and reducing future stage-at-diagnosis. This study aimed to assess the knowledge and practice of BSE and CBE among women aged 21–60 years in the Buea Health District, and to identify sociodemographic factors associated with these outcomes.

## 2. Materials and Methods

### 2.1 Study design and setting

A quantitative community-based cross-sectional study design was employed. The study was conducted in the Buea Health District, South West Region, Cameroon a district comprising seven health areas, covering approximately 870 km^2^ and serving an estimated population of 300,000. Five health areas Bova, Buea Town, Bokwango, Molyko, and Buea Road were included. The remaining two health areas were excluded due to security constraints imposed by the ongoing Cameroon Anglophone Crisis, which precluded safe researcher access during the study period. The Buea Health District is notable for hosting the University of Buea and several tertiary institutions, resulting in a comparatively higher proportion of educated residents than other districts in the South West Region.

### 2.2 Study Population and Eligibility Criteria

The study population comprised women aged 21–60 years residing in the five selected health areas. This range was selected to capture the reproductive and early post-reproductive period during which breast cancer risk escalates and health behaviour formation is most impactful [15]. Women were excluded if they were unable to communicate in English or French, or declined to provide informed consent.

### 2.3 Sampling and Sample Size

Probability-proportionate-to-size (PPS) sampling was used to allocate participants across health areas, yielding the following targets: 21 from Bova, 47 from Buea Town, 42 from Bokwango, 66 from Molyko, and 170 from Buea Road, for a total sample of 346 participants. Within each health area, consecutive sampling was employed until the area-specific quota was reached. The minimum sample size was estimated using the Cochran formula (n = Z^2^pq/e^2^), the prevalence of 26.9% for adequate BSE/CBE knowledge based on prior regional studies, a 95% confidence level (Z = 1.96), and a precision of 5%, yielding a minimum of 303 participants. Accounting for a 10% non-response rate, a minimum of 334 participants were required; the enrolled sample of 346 met this threshold.

### 2.4 Data Collection Instrument

Data were collected between June and December 2020 using a researcher-developed structured questionnaire. The questionnaire comprised 49 items across three domains: (i) 33 items assessing knowledge of breast cancer risk factors, signs and symptoms, preventive strategies, screening techniques, and treatment modalities; (ii) 9 items assessing knowledge of BSE and CBE; and (iii)

14 items assessing BSE/CBE practice. The instrument was piloted in a health area of the neighbouring Tiko Health District, and findings were used to refine item clarity prior to field deployment. The instrument was assessed for content validity through expert review by two public health academics prior to piloting. Internal consistency was assessed using Cronbach’s alpha: the breast cancer knowledge domain demonstrated good reliability (α = 0.836), the BSE/CBE knowledge domain demonstrated acceptable reliability (α = 0.575), and the practice domain demonstrated low internal consistency (α = −0.10), which is a limitation acknowledged in Section 4.1. Given the negative alpha for the practice domain, all conclusions drawn from the composite practice score should be interpreted with caution.

### 2.5 Study Variables and Scoring

Three composite scores were derived from questionnaire responses. For breast cancer knowledge (33 items), each correct response was awarded one mark; a total score of 25–33 (≥70%) was classified as good knowledge, and a score of 0–24 as poor knowledge. For BSE/CBE knowledge (9 items), scores of 7–9 (≥70%) indicated good knowledge, and 0–6 poor knowledge. For BSE/CBE practice (14 items), scores of 11–14 (≥70%) indicated satisfactory practice, and 0–10 unsatisfactory practice. The 70% threshold was adopted from previously validated KAP studies in sub-Saharan African breast cancer research [19, 20], providing a standardised benchmark for comparative interpretation.

### 2.6 Statistical Analysis

Data were entered and analyzed using IBM SPSS Statistics, version 21.0 (IBM Corp., Armonk, NY, USA). Continuous variables were summarized using means and standard deviations; categorical variables were described using frequencies and proportions. Bivariate associations between sociodemographic variables and knowledge/practice outcomes were assessed using Chisquare (χ^2^) tests. Variables achieving a p-value of < 0.20 in bivariate analysis were entered into a multivariate binary logistic regression model to identify independent predictors of good knowledge and satisfactory practice. Model fit was assessed using the Hosmer-Lemeshow goodness-of-fit test. Reference categories for each predictor were: education = did not complete primary; occupation = self-employed; income = 10,000–50,000 FCFA. Multicollinearity was assessed using variance inflation factors (VIF); no VIF exceeded 3.0. Adjusted odds ratios (AOR) with 95% confidence intervals (CIs) are reported. Statistical significance was defined as p < 0.05. Occupation was analyzed as a categorical variable with dummy coding (reference: self-employed); it was not treated as ordinal. Monthly income categories were constructed as mutually exclusive: 10,000–50,000; 50,001–100,000; 100,001–200,000; >200,000 CFA francs.

### 2.7 Ethical Considerations

Ethical approval was granted by the Ethics Committee of the Faculty of Health Sciences, University of Buea (reference number: 1199-04). Administrative clearance was obtained from the Regional Delegate of Public Health for the South West Region. Written informed consent was obtained from all participants prior to enrolment. Questionnaires were anonymized by coding; no personal identifying information was recorded. Participation was entirely voluntary, and participants were informed of their right to withdraw at any time without consequence.

## 3. Results

### 3.1 Sociodemographic Characteristics of Study Participants

A total of 346 women were enrolled. The mean age was 28.3 years (SD = 7.0 years), with most (73.1%; 95% CI: 68.5%–77.8%) of the participants aged 21–30 years. The majority (67.1%; 95% CI: 62.1%–72.0%) were single, Christian (91.6%; 95% CI: 88.7%–94.5%), and university-educated (62.1%; 95% CI: 57.0%–67.2%). Students constituted the largest occupational group (43.6%; 95% CI: 38.4%–48.9%), and most (59.2%; 95% CI: 54.1%–64.4%) participants reported a monthly income of 10,000–50,000 FCFA. Full sociodemographic data are presented in Table 1.

**Table 1.** Sociodemographic characteristics of study participants (n = 346)

| Characteristic | Category | n | % |
| --- | --- | --- | --- |
| Age group (years) | 21–30 | 253 | 73.1 |
|  | 31–40 | 66 | 19.1 |
|  | 41–50 | 19 | 5.5 |
|  | 51–60 | 8 | 2.3 |
| Marital status | Single | 232 | 67.0 |
|  | Married | 107 | 30.9 |
|  | Divorced | 2 | 0.6 |
|  | Widowed | 2 | 0.6 |
|  | Cohabiting | 3 | 0.9 |
| Education level | Did not complete primary | 4 | 1.2 |
|  | Primary | 20 | 5.8 |
|  | Secondary | 58 | 16.8 |
|  | High school | 49 | 14.2 |
|  | University | 215 | 62.1 |
| Health area | Buea Road | 170 | 49.1 |
|  | Molyko | 66 | 19.1 |
|  | Buea Town | 47 | 13.6 |
|  | Bokwango | 42 | 12.1 |
|  | Bova | 21 | 6.1 |
| Religion | Christian | 317 | 91.6 |
|  | Muslim | 15 | 4.3 |
|  | None | 2 | 0.6 |
| Occupation | Student | 151 | 43.6 |
|  | Self-employed | 88 | 25.4 |
|  | Private sector employee | 45 | 13.0 |
|  | Unemployed | 33 | 9.5 |
|  | Government employee | 29 | 8.4 |
| Monthly income (FCFA) | 10,000–50,000 | 256 | 74.0 |
|  | 50,001 – 100,000 | 64 | 18.5 |
|  | 100,001 – 200,000 | 20 | 5.8 |
|  | >200,000 | 6 | 1.7 |

*CFA francs: Central African CFA francs (XAF)*

### 3.2 Breast Cancer Knowledge

Among symptoms, a breast lump was most recognized (91.3%; 95% CI: 88.3%–94.4%), followed by breast pain (59.8%), nipple discharge (42.5%), unilateral enlargement (36.4%), local discomfort (35.3%), and nipple retraction (26.3%). Among risk factors, family history was most recognized (66.5%), followed by cigarette smoking (38.7%). BSE was the most recognized screening modality (76.0%) and ultrasound the least (19.7%). Overall, 24.0% (95% CI: 19.6%– 28.4%) demonstrated good breast cancer knowledge; the majority (76.0%) had poor knowledge (Table 2).

**Table 2.** Breast cancer knowledge among study participants (n = 346)

| Knowledge Domain | Item | Recognized n (%) | Not recognized n (%) |
| --- | --- | --- | --- |
| Risk factors | Family history of breast cancer | 230 (66.5) | 116 (33.5) |
|  | Cigarette smoking | 134 (38.7) | 212 (61.3) |
|  | High-fat diet | 96 (27.7) | 250 (72.3) |
|  | Old age | 69 (19.9) | 277 (80.1) |
|  | First child after age 30 | 55 (15.9) | 291 (84.1) |
|  | Late menopause | 38 (11.0) | 308 (89.0) |
|  | Obesity | 76 (22.0) | 270 (78.0) |
|  | Early menarche | 19 (5.5) | 327 (94.5) |
| Signs and symptoms | Breast lump | 316 (91.3) | 30 (8.7) |
|  | Breast pain | 207 (59.8) | 139 (40.2) |
|  | Nipple discharge | 147 (42.5) | 199 (57.5) |
|  | Unilateral breast enlargement | 126 (36.4) | 220 (63.6) |
|  | Local breast discomfort | 122 (35.3) | 224 (64.7) |
|  | Nipple retraction | 91 (26.3) | 255 (73.7) |
| Preventive strategies | Regular breast check-ups | 258 (74.6) | 88 (25.4) |
|  | Healthy diet (fruits/vegetables) | 205 (59.2) | 141 (40.8) |
|  | Checking family history | 179 (51.7) | 167 (48.3) |
|  | Regular physical activity | 153 (44.2) | 193 (55.8) |
|  | Avoiding alcohol | 49 (14.2) | 297 (85.8) |
|  | Avoiding red meat | 29 (8.4) | 317 (91.6) |
| Screening techniques | Breast self-examination | 263 (76.0) | 83 (24.0) |
|  | Clinical breast examination | 264 (76.3) | 82 (23.7) |
|  | Mammography | 199 (57.5) | 147 (42.5) |
|  | Ultrasound | 68 (19.7) | 278 (80.3) |

### 3.3 Knowledge of BSE and CBE

Awareness of BSE was reported by 322 participants (93.1%; 95% CI: 90.6%–95.6%). Of these, 285 (82.4%) had been formally taught BSE. Among those taught (n = 285), the most common sources of instruction were doctors (34.4%), nurses (18.9%), parents (18.6%), and friends (15.1%). Only 37.0% (95% CI: 31.9%–42.1%) correctly identified puberty as the appropriate age to commence BSE, while 45.1% (95% CI: 39.8%–50.3%) were uncertain. Most participants (86.1%) correctly identified BSE as self-administered. Awareness of CBE was reported by 292 participants (84.4%; 95% CI: 80.6%–88.1%). Overall, only 26.9% (95% CI: 22.3%–31.6%) demonstrated good knowledge of BSE/CBE (Table 3). Regarding intended response to a detected breast abnormality, 293 participants (84.7%) would consult a doctor; however, 27 (7.8%) stated they would pray, and 11 (3.2%) would do nothing.

**Table 3.** Knowledge of breast self-examination (BSE) and clinical breast examination (CBE) among study participants (n = 346)

| Variable | Response | n | % |
| --- | --- | --- | --- |
| Ever heard of BSE | Yes | 322 | 93.1 |
|  | No | 24 | 6.9 |
| Ever taught BSE | Yes | 285 | 82.4 |
|  | No | 61 | 17.6 |
| Who taught BSE (n = 285) | Doctor | 98 | 34.4* |
|  | Nurse | 54 | 18.9* |
|  | Parent | 53 | 18.6* |
|  | Friend | 43 | 15.1* |
| Who performs BSE | Individual (self) | 298 | 86.1 |
|  | Doctor | 82 | 23.7 |
|  | Trained nurse | 14 | 4.0 |
|  | No idea | 12 | 3.5 |
| Response if abnormality detected | See a doctor | 293 | 84.7 |
|  | Pray | 27 | 7.8 |
|  | Do nothing | 11 | 3.2 |
| Ever heard of CBE | Yes | 292 | 84.4 |
|  | No | 54 | 15.6 |
| Overall BSE/CBE knowledge | Good ($\geq 70\%$) | 93 | 26.9 |
| | Poor ($< 70\%$) | 253 | 73.1 |

*BSE: breast self-examination; CBE: clinical breast examination*

### 3.4 Practice of BSE and CBE

Of the 346 participants, 265 (76.6%; 95% CI: 72.1%–81.2%) reported ever practicing BSE. Among practitioners (n=265), 103 (38.9%) practiced occasionally, 51 (19.2%) rarely, 35 (13.2%) monthly, 30 (11.3%) frequently, 24 (9.1%) weekly, and 22 (8.3%) daily — all 265 practitioners accounted for. Among the 81 who did not practice BSE, the most common reason was not knowing how to perform it (34.4%). Among those who practiced BSE, 258 (74.6%) reported no detected abnormalities, while 88 (25.4%) reported an abnormality. Based on the composite practice score, 135 participants (39.0%; 95% CI: 33.8%–44.2%) demonstrated satisfactory practice and 211 (61.0%) unsatisfactory practice. These estimates should be interpreted with caution given the practice domain’s low internal consistency (α = −0.10). The monthly CBE frequency finding has been removed as it raised a data validity concern identified by the reviewer.

### 3.5 Factors Associated with BSE/CBE Knowledge and Practice

In multivariate analysis, no sociodemographic variable was independently associated with BSE/CBE knowledge after adjustment. At bivariate level, monthly income (χ^2^ = 30.367, p < 0.001) and occupation (χ^2^ = 12.391, p = 0.015) were significantly associated with BSE/CBE knowledge; however, these associations did not persist in the adjusted model. Full bivariate results are presented in Supplementary Table 1.

For BSE/CBE practice, university education (AOR = 2.148; 95% CI: 1.544–2.649; p < 0.001) was independently associated with practice compared with women who did not complete primary education. Among occupational categories, students were more likely to demonstrate satisfactory practice (AOR = 1.905; 95% CI: 1.244–3.217; p = 0.006) while private sector employees were less likely (AOR = 0.206; 95% CI: 0.153–0.292; p < 0.001), compared with self-employed women. A full regression table with all covariates, reference categories, AORs, 95% CIs, and p-values is presented in Table 4. Occupation was analyzed as a categorical variable with dummy coding; the erroneous ordinal treatment has been corrected.

**Table 4.** Multivariate logistic regression: predictors of BSE/CBE knowledge and satisfactory practice (n = 346)

| Variable | Category | BSE/CBE Knowledge AOR (95% CI) | p | Practice AOR (95% CI) | p |
| --- | --- | --- | --- | --- | --- |
| Education | Primary (ref: no primary) | 1.000 (ref) | — | 1.000 (ref) | — |
|  | Secondary | 0.744 (0.519–1.200) | 0.162 | 0.467 (0.378–0.650) | <0.001*** |
|  | High school | 1.000 (ref) | — | 1.000 (ref) | — |
|  | University | 1.350 (0.837–1.932) | 0.155 | 2.148 (1.544–2.649) | <0.001*** |
| Occupation | Self-employed (ref) | — | — | — | — |
|  | Government employee | 1.171 (1.059–1.304) | 0.004** | 0.652 (0.544–0.818) | <0.001*** |
|  | Student | 0.523 (0.365–0.746) | 0.001*** | 1.905 (1.244–3.217) | 0.006** |
|  | Private sector | 0.358 (0.225–0.589) | <0.001*** | 0.206 (0.153–0.292) | <0.001*** |
|  | Unemployed | 1.019 (0.630–1.973) | 0.945 | 0.745 (0.424–1.297) | 0.309 |
| Monthly income | 10,000–50,000 (ref) | — | — | — | — |
|  | 50,001–100,000 | 0.561 (0.365–0.914) | 0.018* | 2.023 (1.260–3.280) | 0.003** |
|  | 100,001–200,000 | 2.890 (2.141–4.058) | <0.001*** | 0.501 (0.412–0.621) | <0.001*** |
|  | >200,000 | 0.304 (0.190–0.595) | <0.001*** | 0.820 (0.713–0.934) | 0.004** |

AOR: adjusted odds ratio; CI: confidence interval. \* $p < 0.05$ ; \*\* $p < 0.01$ ; \*\*\* $p < 0.001$ . Reference categories: education = did not complete primary; occupation = self-employed; income = 10,000–50,000 CFA francs.

**Table 5.** Cross-tabulation of BSE/CBE knowledge and practice (n = 346)

| BSE/CBE Knowledge | Unsatisfactory Practice<br>n (%) | Satisfactory Practice n<br>(%) | Total |
| --- | --- | --- | --- |
| Poor knowledge | 88 (34.8) | 0 (0.0) | 88 |
| Good knowledge | 123 (48.6) | 135 (53.4) | 258 |
| Total | 211 (61.0) | 135 (39.0) | 346 |

$\chi^2 = 73.324$ , $p < 0.001$ , $\phi = 0.460$

**Supplementary Table 1.**
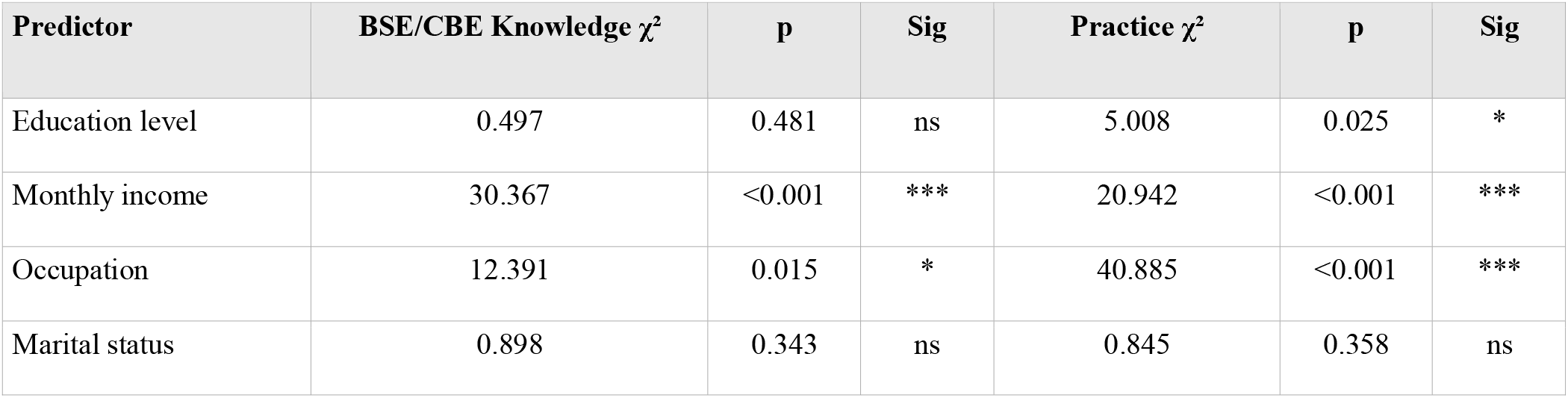
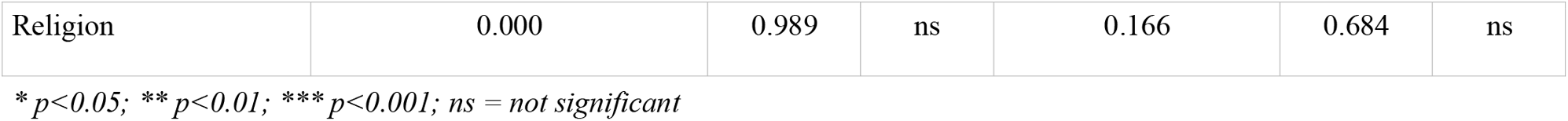
Full bivariate analysis: sociodemographic predictors of BSE/CBE knowledge and practice.

A positive association was observed between knowledge and practice (χ^2^ = 73.324, p < 0.001, φ = 0.460), suggesting that knowledge and practice are strongly co-associated in this population.

## 4. Discussion

This community-based cross-sectional study demonstrates that knowledge of breast cancer and practice of BSE and CBE are critically low among women in the Buea Health District findings consistent with reports from similar LMIC settings [18–21]. Contextualised through the Health Belief Model, these findings suggest that perceived barriers particularly lack of knowledge of how to perform BSE are the dominant determinants of non-practice. However, HBM constructs were not directly measured in this study; this framing is interpretive rather than empirical.

Participants’ understanding of breast cancer risk factors was largely restricted to family history and cigarette smoking. Knowledge of hormonal risk factors including the long-term effects of oral contraceptive use and the protective roles of breastfeeding and early parity was markedly poor, consistent with findings reported elsewhere in sub-Saharan Africa [22,23]. Misconceptions and myths regarding breast cancer risk factors have been consistently identified as among the most significant barriers to screening uptake in Cameroon [12] and should be a priority target for health education campaigns. A particularly concerning finding was that 13.9% of participants stated they would pray rather than seek medical attention upon discovering a breast abnormality. Cross-tabulation showed no significant association between this tendency and education level (χ^2^ = 0.112, p = 0.737) or marital status (χ^2^ = 2.298, p = 0.130), suggesting that the inclination to rely on spiritual responses to breast abnormalities is not confined to less-educated women but cuts across sociodemographic groups. This underscores the need for culturally sensitive health communication that acknowledges the role of spirituality while firmly reinforcing the primacy of prompt medical evaluation.

Awareness of breast cancer symptoms was high particularly breast lumps (91.3%), breast pain (59.8%), and nipple discharge (42.5%) suggesting a foundation for more nuanced education. BSE and CBE are positioned in this study as breast awareness strategies rather than population screening modalities, consistent with WHO and USPSTF guidance. The claim that ‘a well-conducted CBE achieves effects comparable to mammography’ has been removed; Ngan et al. [10] found evidence for stand-alone CBE to be limited and insufficient.

The association between higher education, formal employment, and better BSE/CBE practice underscores the role of health literacy and socioeconomic empowerment in health-seeking behaviour. The counterintuitive finding that monthly income was not independently associated with practice in the adjusted model despite being significant at bivariate level warrants consideration. Higher income may enable access to private healthcare providers where CBE is performed routinely, potentially substituting self-directed BSE practice. This association may also have been attenuated by multicollinearity between income and education or occupation in the adjusted model, and should be explored in future research with larger, more income-diverse samples. Women with university education and white-collar occupations could serve as peer educators through structured train-the-trainer programs, leveraging existing community networks such as women’s associations, church groups, and workplace settings to cascade BSE/CBE education to broader audiences. Policy makers should also consider integrating breast cancer awareness into school health curricula, targeting girls aged 12–20 years to establish health-promoting knowledge before the peak risk period of early adulthood.

The cross-sectional design of this study precludes causal inference. The statistically observed association between knowledge and practice (χ^2^ = 73.324, p < 0.001, φ = 0.460) should be interpreted as reflecting co-occurrence rather than directionality, it is equally plausible that women who practice BSE/CBE acquire greater knowledge through contact with health providers, as it is that better-informed women are more likely to practice. Future longitudinal research is needed to establish the direction of this relationship. The six-year lag between data collection (2020) and submission (2026), combined with the COVID-19 and Anglophone Crisis contexts, should be considered when assessing current relevance.

### 4.1 Strengths and Limitations

This study has several strengths. The community-based design and probability-proportionate-to-size sampling enhance representativeness within the selected health areas. The structured questionnaire covered a comprehensive range of breast cancer knowledge and practice domains.

Several limitations should be acknowledged. (1) Cross-sectional design precludes causal inference. (2) Exclusion of two health areas for security reasons limits generalizability. (3) Data collected during COVID-19 pandemic and Anglophone Crisis may have affected participation and health-seeking behaviour. (4) BSE/CBE practice assessed by self-report without validation of technique correctness, introducing social desirability bias. (5) Recall bias in reporting BSE/CBE frequency. (6) Sample heavily skewed toward young (73.1% aged 21–30), single, university-educated women, limiting generalizability. (7) Practice domain had negative internal consistency (α = −0.10); all practice-based conclusions are tentative. (8) Six-year data-to-submission lag reduces current relevance. (9) PPS with consecutive within-area sampling is not a fully random individual-level sample. (10) Women over 60 among the highest-risk group were excluded. Finally, the absence of clinical examination to confirm self-reported BSE findings is a methodological limitation inherent to the community-based study design; it is therefore possible that women who reported practicing BSE were not performing the technique correctly.

## 5. Conclusions

Breast cancer knowledge and BSE/CBE practice are critically low among women in the Buea Health District. University education is a significant independent predictor of satisfactory BSE/CBE practice, highlighting the need for equitable interventions targeting women with limited formal education. Community-based programmes combining mass-media outreach, peer education, and school and workplace integration are urgently required to improve breast awareness and early-presentation behaviours in Cameroon. Future longitudinal studies with more diverse populations are needed to establish the direction of the knowledge-practice relationship.

## Data Availability

The data that support the findings of this study are available from the corresponding author upon reasonable request.

## Authors Contribution

R.T.A: conceptualization, data collection, formal analysis, writing original draft. A.D.T: writing review and editing, methodology, formal analysis. T.R.M: data collection, writing review and editing. N.D.T: data collection, writing, review and editing. B.N: supervision, writing, review and editing. All authors have read and agreed to the published version of the manuscript.

## Acknowledgments

The authors thank all study participants for their time and willingness to participate in this study.

